# Polygenic Parkinson’s disease genetic risk score as risk modifier of parkinsonism in Gaucher disease

**DOI:** 10.1101/2022.12.19.22280175

**Authors:** Cornelis Blauwendraat, Nahid Tayebi, Elizabeth Geena Woo, Grisel Lopez, Luca Fierro, Marco Toffoli, Naomi Limbachiya, Derralynn Hughes, Vanessa Pitz, Dhairya Patel, Dan Vitale, Mathew J. Koretsky, Dena Hernandez, Raquel Real, Roy N. Alcalay, Mike A Nalls, Huw R Morris, Anthony H. V. Schapira, Manisha Balwani, Ellen Sidransky

## Abstract

**Background:** Bi-allelic pathogenic variants in *GBA1* are the cause of Gaucher disease (GD1), a lysosomal storage disorder resulting from deficient glucocerebrosidase. Heterozygous *GBA1* variants are also a common genetic risk factor for Parkinson’s disease (PD). GD manifests with considerable clinical heterogeneity and is also associated with an increased risk of PD.

**Objective:** To investigate the contribution of PD risk variants to risk of PD in patients with GD1.

**Methods:** We studied 225 patients with GD1, including 199 without PD and 26 with PD. All cases were genotyped and the genetic data was imputed using common pipelines.

**Results:** On average, patients with GD1 with PD have a significantly higher PD genetic risk score than those without PD (P=0.021).

**Conclusions:** Our results indicate that variants included in the PD genetic risk score were more frequent in patients with GD1 who developed PD, suggesting that common risk variants may affect underlying biological pathways.

Supplemental data <u>here</u>

## Introduction

Pathogenic variants in *GBA1*, the gene encoding the enzyme glucocerebrosidase (GCase) are associated with the recessively inherited lysosomal storage disorder Gaucher disease (GD). *GBA1* mutations are also the most common known genetic risk factor for Parkinson’s disease (PD) and dementia with Lewy bodies ^1,2^. Traditionally, GD is divided into three clinical types based on the severity of the disease and the degree of neurological involvement: type 1 is non-neuronopathic, type 2 is the acute neuronopathic form presenting in infancy, and type 3 is chronic neuronopathic. There is a wide spectrum of disease manifestations and disease severity, even within the different types ^3^. Approximately 600 GD causing *GBA1* variants have been described, of which the vast majority in the heterozygous state show also increased risk for PD. Certain *GBA1* variants (including p.E365K and p.T408M ^4,5^) are considered PD risk factors and affect PD progression but do not cause GD, even in homozygotes, although they may produce a modest reduction in GCase activity ^6^. Genotype/phenotype correlations for GD are somewhat limited, although some pathogenic variants are more likely to be associated with a specific type of GD. For example, the common N370S (p.N409S) mutation is exclusively seen in GD type 1 (GD1). By definition, patients with GD1 do not have CNS involvement although they are at increased risk for developing PD and DLB. However, the great majority of patients with GD do not exhibit parkinsonism, even as they age. While there are no reliable large scale studies, estimates are that 8-12% of patients with GD1 at age 80 report PD symptoms ^7,8^, indicating that a majority of patients with GD1 escape PD. Thus, patients with the lysosomal storage disorder GD1 provide a unique cohort for exploring other secondary (potentially genetic) factors predisposing to or protecting from the development of PD ^5^. While these patients have a major deficiency of GCase and often multisystem involvement, it is essential to understand why only a subset develop PD. Here we performed genome-wide genotyping on a relatively large cohort of GD1 subjects and assess the contribution of common variants included in the most recent PD risk score ^9^ to PD risk in patients with GD1.

## Subjects and Methods

### Cohort information

GD1 cases were collected from three different sites: NHGRI, Mt Sinai and UCL/Royal Free London Hospital NHS Foundation Trust. The NHGRI GD dataset was collected over the past two decades and includes patients with a verified diagnosis of Gaucher disease evaluated at the Clinical Center of the National Institutes of Health under an IRB approved clinical protocol. *GBA1* genotyping, performed on all individuals, was done by Sanger sequencing all exons of the gene. Patients were evaluated by a movement disorder specialist for signs of PD. A subset of the cohort was followed longitudinally for up to 15 years. Mount Sinai cases were collected via an IRB-approved study beginning January 2019. Patients were evaluated at the Lysosomal Storage Disease program, had a confirmed diagnosis of GD and were screened for PD, with the diagnosis confirmed by a movement disorder specialist. UCL GD cases were collected through UK Lysosomal Storage Diseases Centres, clinically assessed and all *GBA1* exons genotyped, through a project approved by the local ethics (IRB) committee. PD symptoms and diagnosis were all confirmed by a movement disorder specialist. Information on age of onset of motor symptoms for GD1 with PD and age at last exam for GD1 was also collected. Additional PD and control data were obtained from the Global Parkinson’s Genetics Program (GP2) (release 1) which is genotyped on the same genotyping array, facilitating comparison with GD1 and GD1-PD data ^10^. More cohort details are provided in Supplemental Table 1.

### Genotyping of DNA and data analysis

Genotyping was performed at NIH/LNG and UCL Genomics centers using the new Global Diversity Array (GDA) with NeuroBooster content (https://github.com/GP2code/Neuro_Booster_Array). Genetic data was cleaned and imputed using standard GP2 pipelines and protocols (https://github.com/GP2code/). In brief, sample level quality control steps was performed using PLINK (1.9 and 2) and includes genotype missingness (<0.02), genetic sex confirmation, duplicate check and relatedness confirmation. We excluded a random sample if relatedness higher than 0.2 was identified, although if one of the related samples was a GD1-PD case, that sample was retained. Variants were filtered for missingness and other standard parameters and imputed using TOPMed Imputation Server ^11,12^. Ancestry was defined using 1000 Genomes and an Ashkenazi Jewish (AJ) reference population ^13,14^. Principal component (PC) analysis was performed using PLINK, and when plotting, genetic ancestries were color-coded (Supplementary Fiigure 1). Additional PCs were generated per ancestry and included in subsequent analyses as covariates together with age of onset for PD cases and age of last exam for non-PD subjects. Age was missing for 27 patients and these were imputed to the mean. The PD genetic risk score was calculated using imputed data. We used weights from known GWAS variants from Nalls et al 2019, excluding the full *GBA1* region, and two additional variants were filtered out due to high missingness >5% (chr10:119776815:G:A and chr19:2341049:C:T) leaving 85 variants. The genetic risk score was normalized based on GP2 controls from each population (AJ and EUR), with one unit of change representing a single standard deviation increase in the control risk score. Case-control analysis was performed using a logistic regression using biological sex and five PCs as covariates. Meta-analysis was performed across ancestries using a conservative random effects model. All calculations and figures were made with R (3.6.1 or 4.2.0) using packages ggplot2, metafor, rmeta. All code used can be found at https://github.com/neurogenetics/Gaucher_PD_GRS_modifiers

## Results

To investigate genetic modifiers of risk of PD symptoms in patients with GD1, we genotyped three GD1 cohorts totaling 266 GD1 cases, of which 27 were also diagnosed with PD. Unfortunately, the analysis was only possible in the European (EUR) and Ashkenazi Jewish subgroups due to insufficient GD1-PD cases for analysis in the other ancestral groups. After further sample level quality control, this resulted in 26 GD1-PD cases (18 AJ and 8 EUR) and 199 GD1 cases without PD (134 AJ and 65 EUR). As additional data for comparison, we included PD cases (335 AJ and 2050 EUR) and controls (109 AJ and 933 EUR) from GP2 (Supplemental Table 2).

Using the imputed data we calculated the PD genetic risk score for each participant and normalized the values based on the GP2 controls of each ancestry. The genetic risk score is the cumulative dosage of risk alleles with each SNP’s contribution weighted by its identified GWAS effect estimate as a means of predicting risk. First, we compared the genetic risk score for PD vs controls per ancestry and meta-analyzed results. Here we identified a significant difference (P=1.18E-24) with a highly similar effect size (OR=1.575, CI=1.444-1.717, I2=0) as previously described ^9^ (Figure 1A, B, C, Supplemental Table 3). Next, we performed a similar analysis using GD1-PD cases and GD1 without PD. We identified a significant effect (P=0.0168, OR=1.687, CI=1.099-2.589) between GD1-PD cases and GD1 without PD (Figure 1A, C and 1D, Supplemental Table 4). No significant heterogeneity of effect was detected (I2=0.0) and the effect size was remarkably similar to the PD vs control. Of note, two AJ individuals were identified as carriers of *LRRK2* p.G2019S, one with GD1-PD and one GD1 alone.

**Figure 1:**
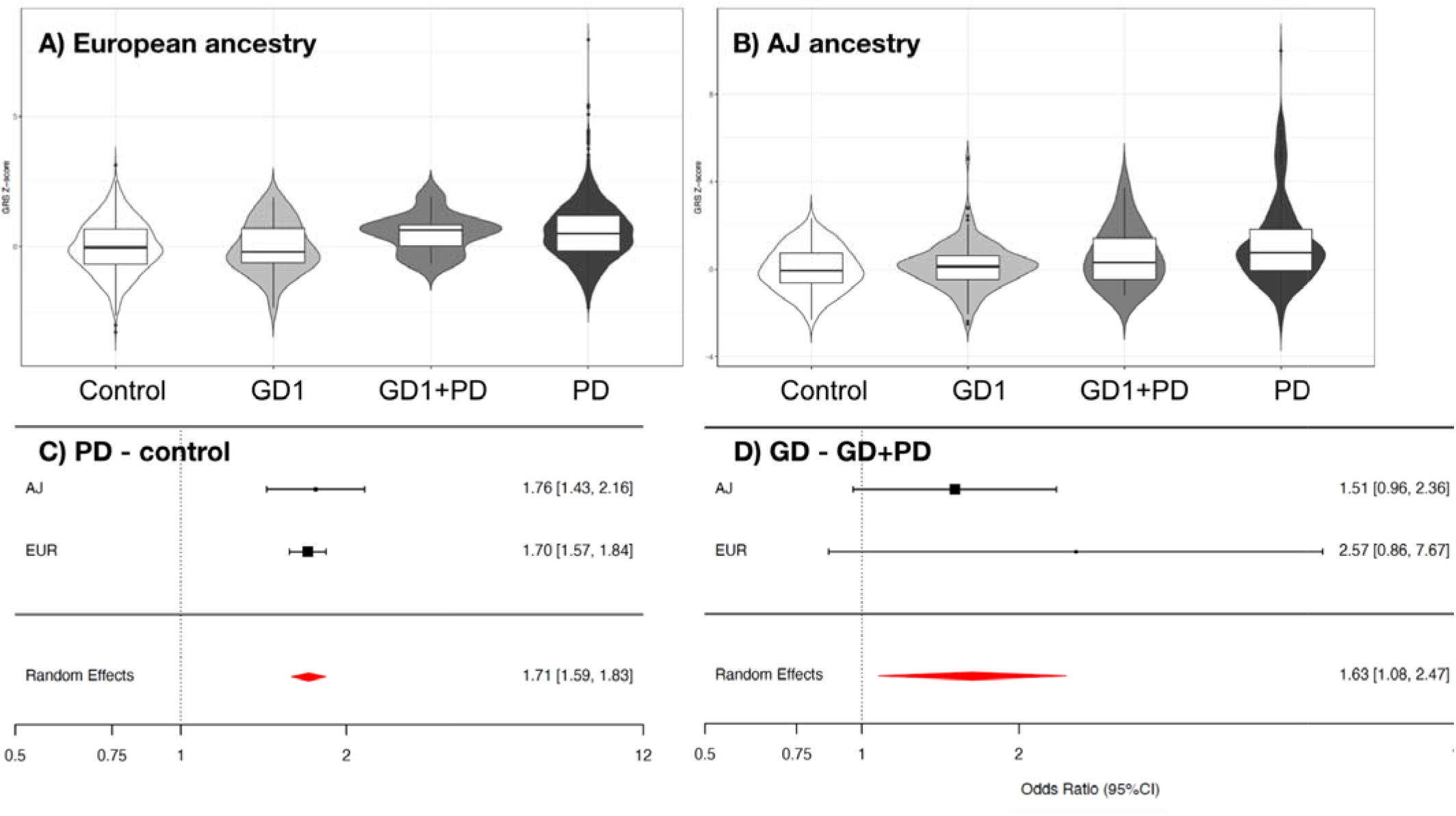
Parkinson’s disease (PD) genetic risk score comparisons across cohorts and ancestries. A) Violin plots of genetic risk score of European (EUR) ancestry control, Gaucher disease type 1 (GD1) without PD, GD1 with PD and PD cases show a higher risk score (normalized Z-score) in both GD1+PD and PD vs control and GD1 respectively. B) Identical plots for subjects with Ashkenazi Jewish (AJ) ancestry. C) Forest plot of PD - control risk score analysis of EUR and AJ ancestries shows a significant effect. D) Forest plot of GD1 without PD - GD1+PD risk score analysis of EUR and AJ ancestries shows a significant effect. Bracket end lines denote 95% CIs of the study specific estimates, with squares for effect estimates (odds ratios), and the size of these squares is proportionate to sample size. The red diamonds denote effect and 95% CI of the meta-analyzed results on the odds ratio scale.

## Discussion

Current estimates of patients with GD1 that report PD symptoms by age 80 range from 8-12% which is significantly higher than the general risk for PD, estimated at 2% ^15^. It is clear that the majority of patients with GD1 do not develop PD symptoms, suggesting that there are other modifiers of risk. Here we assessed potential genetic modifiers by investigating the PD genetic risk score and determined that the PD genetic score contributes to the risk of PD symptoms in patients with GD1. Interestingly, we previously identified a similar contribution of the PD genetic risk score in *LRRK2* p.G2019S and *GBA1* heterozygous carriers ^16,17^.

Other lysosomal storage disorder genes have also been implicated in PD by rare variant analysis ^18^ and several lysosomal pathways are associated with PD ^9^, emphasizing the importance of the lysosome in PD pathogenesis. Currently, the most prevalent hypothesis linking *GBA1* mutations to PD posits a connection between α-synuclein aggregation and reduced GCase activity ^19,20^. Our results suggest that the common risk variant component of PD contributes to this complex interplay between *GBA1* and α-synuclein aggregation and can increase the risk of developing PD even in GD1 cases, which are already at higher risk of developing PD.

Naturally this study has some limitations. The number of patients with both GD1 and PD included is relatively small (n=28). Since GD1 is a very rare disease with a prevalence of approximately 1 in 100,000 in the general population ^21^ and with an estimate of ∼10% developing PD symptoms, obtaining more participants is challenging. Due to our relatively small numbers, we also could not investigate the effect of specific *GBA1* variants on PD risk in GD1 patients, as well as on the onset and severity of symptoms, although this is an area of interest requiring further study. All three sites conducted longitudinal assessments over the past years/decades, with evaluations not specifically designed for this study. Therefore, harmonized clinical data regarding specific clinical measurements such as motor signs or prodromal non-motor symptoms of PD are not available. We also lack ancestral diversity due to both the low number of participants and the demographics of the referral centers participating. We are planning to expand this cohort and welcome future collaborations. Furthermore, with limited power to perform single variant testing, we could not define GD1 PD specific risk estimates, and used the previously established PD risk estimates from previous GWAS. Additionally, we could not identify which specific PD GWAS loci contribute to the PD risk in the participating patients with GD1.

In summary, here, for the first time, we show that a PD genetic risk score is a risk modifier of PD symptoms in patients with GD1, which has potentially interesting biological implications. Together with our previous work ^16^, our findings provide clear evidence of substantial overlap in the genetic risk affecting *GBA1*-associated PD and typical PD. Thus, identifying the specific factors impacting risk in *GBA1*-PD may yield shared mechanisms underlying other forms of PD. Future studies should carefully phenotype larger cohorts of GD1 patients to accurately estimate age-specific penetrance of PD and identify other modifiers.

## Data Availability

All data produced in the present work are contained in the manuscript.

https://github.com/GP2code/Neuro_Booster_Array

https://amp-pd.org/

## Acknowledgments

This work was supported in part by the Intramural Research Program of the National Human Genome Research Institute (NHGRI), the Center for Alzheimer’s and Related Dementias, within the Intramural Research Program of the National Institute on Aging and the National Institute of Neurological Disorders and Stroke, National Institutes of Health, Department of Health and Human Services, and the National Institutes of Health and Cure Parkinson’s (UK). Data used in the preparation of this article were obtained from Global Parkinson’s Genetics Program (GP2). GP2 is funded by the Aligning Science Across Parkinson’s (ASAP) initiative and implemented by The Michael J. Fox Foundation for Parkinson’s Research (https://gp2.org). For a complete list of GP2 members see https://gp2.org.

## Author Roles

1. Research project: A. Conception, B. Organization, C. Execution;
2. Cohort generation: A. Cohort recruitment, B. Sample processing, C. Data generation:
3. Statistical Analysis: A. Design, B. Execution, C. Review and Critique;
4. Manuscript Preparation: A. Writing of the first draft, B. Review and Critique;

CB: 1A,B,C, 3A,B,C, 4A

NT: 1B, 2A,B,C, 4B

GW: 2A,B,C, 4B

GL: 2A,B,C, 4B

LF: 2A,B,C, 4B

MT: 2A,B,C, 4B

NL: 2A,B,C, 4B

DH: 2A,B,C, 4B

VP: 3B, 4B

DP: 3B, 4B

DV: 3B,C, 4B

MJK: 3B,C, 4B

DH: 2C, 4B

RR: 2C, 4B

RNA: 2A,B,C, 4B

MAN: 3A,B,C, 4B

HRM: 2C, 4B

AHVS: 2A,B,C, 4B

MB: 2A,B,C, 4B

ES: 1A,B,C, 2A,B,C, 4A

