## Supplemental data for "Polygenic Parkinson’s disease genetic risk score as risk modifier of parkinsonism in Gaucher disease"

**Supplemental Figure and Tables**

Supplemental Figure 1: Principal component analysis plot of all included GD1 participants using GenoTools ancestry predictions (<https://github.com/dvitale199/GenoTools>). AJ=Ashkenazi Jewish, EUR=European, AMR= Latino/American Admixed, AAC= African Admixed and Caribbean, EAS=East Asian

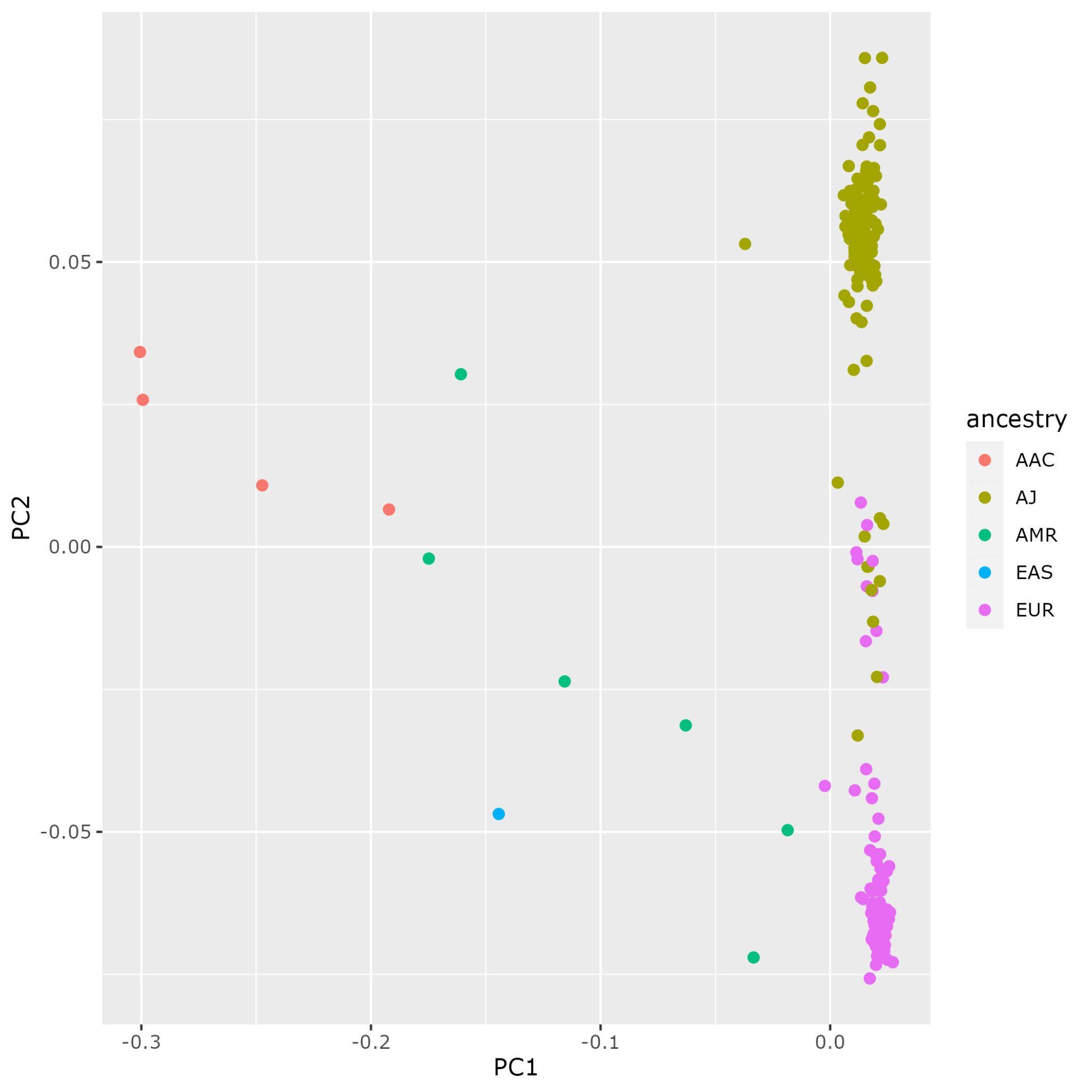

**Supplemental Table 1:** Included cohort overview pre quality control filtering. AJ=Ashkenazi Jewish, EUR=European, AMR= Latino/American Admixed, AAC= African Admixed and Caribbean, EAS=East Asian, GD=Gaucher disease, PD=Parkinson’s disease

| **Cohort** | **Ancestry** | **GD1** | **GD1+PD** | **PD** | **Control** |
| --- | --- | --- | --- | --- | --- |
| NHGRI n=141 | AJ | 66 | 14 | 0 | 0 |
|  | EUR | 44 | 6 | 0 | 0 |
|  | AMR | 5 | 1 | 0 | 0 |
|  | AAC | 4 | 0 | 0 | 0 |
|  | EAS | 1 | 0 | 0 | 0 |
| Mt Sinai n=97 | AJ | 83 | 4 | 0 | 0 |
|  | EUR | 9 | 0 | 0 | 0 |
|  | AMR | 1 | 0 | 0 | 0 |
| UCL n=28 | AJ | 3 | 0 | 0 | 0 |
|  | EUR | 21 | 2 | 0 | 0 |
| GP2 n=3427 | EUR | 0 | 0 | 2050 | 933 |
|  | AJ | 0 | 0 | 335 | 109 |
|  | TOTAL | 239 | 27 | 2385 | 1042 |

**Supplemental Table 2:** Demographic details included GD1 Ashkenazi Jewish (AJ) and European (EUR) in the final analysis after quality control. ^ age not available for 27 individuals, *age of PD onset was not available for 1 subject. GD=Gaucher disease, PD=Parkinson’s disease

| **Cohort** | **GD1** | **GD1+PD** | **Total** |
| --- | --- | --- | --- |
| NHGRI total (AJ/EUR) | 88 (52/36) | 20 (14/6) | 108 |
| Age of last exam (no PD) and age of onset PD Mean (range) | 53.8 (7-97)^ | 51.1 (32-69)* |  |
| male/females | 34/54 | 10/10 |  |
| Mt Sinai total (AJ/EUR) | 87 (79/8) | 4 (4/0) | 91 |
| Age of last exam (no PD) and age of onset PD Mean (range) | 54.2 (26-90) | 51 (43-59) |  |
| male/females | 51/36 | 2/2 |  |
| UCL total  (AJ/EUR) | 24 (3/21) | 2 (0/2) | 26 |
| Age of last exam (no PD) and age of onset PD Mean (range) | 61.8 (27-83) | 58.5 (51-66) |  |
| male/females | 15/9 | 0/2 |  |
| TOTAL | 199 | 26 | 225 |

**Supplemental Table 3:** Statistics of PD case - control analysis

| Ancestry | Beta | OR | CI 95% | SE | Z | P | N_case | N_control |
| --- | --- | --- | --- | --- | --- | --- | --- | --- |
| AJ | 0.5429 | 1.721 | 1.363-2.173 | 0.11897 | 4.563 | 5.04E-06 | 335 | 109 |
| EUR | 0.4398 | 1.552 | 1.414-1.705 | 0.04774 | 9.213 | 3.17E-20 | 2050 | 933 |
| META | 0.4541 | 1.575 | 1.444-1.717 | 0.0443 | 10.2496 | 1.18E-24 | 2385 | 1042 |

**Supplemental Table 4:** Statistics of GD1+PD - GD1 no PD

| Ancestry | Beta | OR | CI 95% | SE | Z | P | N_GD1_PD | N_GD1 |
| --- | --- | --- | --- | --- | --- | --- | --- | --- |
| AJ | 0.4458 | 1.561 | 0.981-2.487 | 0.2374 | 1.878 | 0.0603 | 18 | 134 |
| EUR | 0.9520 | 2.591 | 0.863-7.774 | 0.5606 | 1.698 | 0.0895 | 8 | 65 |
| META | 0.5228 | 1.687 | 1.099-2.589 | 0.2186 | 2.3916 | 0.0168 | 26 | 199 |
